# A reporter virus particle seroneutralization assay for tick-borne encephalitis virus overcomes ELISA limitations

**DOI:** 10.1101/2024.01.23.24301657

**Authors:** Rahel Ackermann-Gäumann, Alexis Dentand, Reto Lienhard, Mohsan Saeed, Margaret R. MacDonald, Alix T Coste, Valeria Cagno

**Affiliations:** Swiss National Reference Centre for Tick-Transmitted Diseases, Switzerland; ADMED Microbiologie, La Chaux-de-Fonds 2300, Switzerland; Lausanne University Hospital, University of Lausanne, Switzerland; Department of Biochemistry & Cell Biology, Boston University Chobanian and Avedisian School of Medicine, Boston University, MA 02118; National Emerging Infectious Diseases Laboratories, Boston University, MA 02118; Laboratory of Virology and Infectious Disease, The Rockefeller University, New York NY 10065

**Keywords:** seroneutralization, TBEV, reporter viral particle, serology

## Abstract

**Background:** Tick-borne encephalitis (TBE) virus is the most common tick-transmitted Orthoflavivirus in Europe. Due to its non-specific symptoms, TBE is primarily diagnosed by ELISA-based detection of specific antibodies in the patient serum. However, cross-reactivity between orthoflaviviruses complicates the diagnosis. Specificity problems may be overcome by serum neutralization assays (SNT), however clinically relevant orthoflaviviruses require handling in biosafety level 3 (BSL-3) and they have highly divergent viral kinetics and cell tropisms.

**Methods:** We present a reporter viral particle (RVP) based SNT in which the infectivity is measured by luminescence and that can be performed under BSL-2 conditions.

**Findings:** The RVP-based SNT for TBEV exhibited a remarkable correlation with the traditional virus-based SNT (R2=0.8614, p<0.0001). Notably, the RVP-based assay demonstrated a sensitivity of 91.7% (95% CI: 87.2-97.1%) and specificity of 100% (95% CI: 79.6-100%). We also tested the cross-reactivity of serum samples in RVP-based assays against other orthoflaviviruses (yellow fever virus, dengue virus type 2, Zika virus, West Nile virus and Japanese encephalitis virus). Interestingly, in 90% of cases where a serum sample had tested TBEV-positive by ELISA but negative by RVP-based SNT, we identified antibodies against other orthoflaviviruses.

**Interpretations:** The RVP-based seroneutralization assay show clinical relevance and broad- applicability.

**Funding:** This study was supported by Bavarian Nordic grant to R.A. and V.C.

**RESEARCH IN CONTEXT:** *Evidence before this study:* ELISA tests for orthoflavivirus serology are the method of choice in all diagnostic laboratories despite the cross-reactivity issues. Although seroneutralization testing (SNT) provides more reliable results, it requires BSL-3 conditions and approximately a week to obtain the results. However, developing tests with a broader applicability could overcome the problem of cross-reactivity of antibodies against flaviviruses could be overcome leading to a more accurate diagnosis and fewer non-useful results. Although alternative serological tests for other orthoflaviviruses have been investigated they have limitations, including lack of uniformity for different orthoflaviviruses, the need for a BSL-3 laboratory to perform them, and results taking 4-5 days. The reporter viral particle system (RVP) we used in this study has been reported for all orthoflaviviruses, except for YFV. However, its applicability has not been tested in comparison to traditional methods with clinical samples.

*Added value of this study:* We tested the RVP system uniformly for different orthoflaviviruses and evaluated the sensitivity and specificity of SNT based on RVP compared to virus-based and to ELISA. Additionally, we found that false positives in ELISA in our clinical samples are frequently related to YFV positive samples.

*Implications of all the available evidence:* This study demonstrates the reliability and broad applicability of implementing RVP-based SNT in a clinical setting. This test can overcome the issues of false positive results from ELISA tests. Additionally, our data suggest that it is important to consider YFV exposure or vaccination anamnesis in patient’s medical history. This is consistent with the phylogenetic similarity between YFV and TBEV if compared to other flaviviruses.

## INTRODUCTION

Tick-borne encephalitis (TBE) caused by the tick-borne encephalitis virus (TBEV; Orthoflavivirus encephalitidis, genus Orthoflavivirus, family *Flaviviridae*), is a major public health problem in large parts of Europe and Asia. In Europe, TBE is endemic in at least 27 countries. TBE is typically caused by an infection involving one of three TBEV subtypes, namely the European, Siberian, and Far Eastern subtypes. In addition, two other subtypes, i.e. the Baikalian and the Himalayan subtype have been described more recently (1). TBEV is primarily transmitted to humans by the bite of an infected Ixodid tick, although transmission via the consumption of unpasteurized milk products from infected goats, sheep, or cows, is also possible (2).

Despite the availability of effective vaccines, disease incidence has significantly increased during the past years (1). TBE may manifest as a disease of variable severity, ranging from subclinical infections to severe courses with neurological involvement and potentially fatal outcomes. Symptomatic disease is typically biphasic when caused by European subtype viruses, including a viremic stage with flu-like symptoms starting about 8 days (range 4–28 days) after the tick bite, an asymptomatic interval of about one week (range 1–33 days), and a second stage with neurological manifestations ranging from mild meningitis to severe encephalitis with or without myelitis and spinal paralysis (1, 3, 4).

TBEV virions are spherical and contain a nucleocapsid surrounded by a lipid bilayer. The nucleocapsid consists of single-stranded positive-sense genomic RNA and the capsid protein (C). The surface of the lipid membrane incorporates an envelope (E) and a membrane (M) glycoprotein (5). In addition to the three structural proteins (C, E, and precursor M [prM]), the viral genome encodes for seven nonstructural proteins (NS1, NS2A, NS2B, NS3, NS4A, NS4B, and NS5).

Since clinical symptoms of TBE are often unspecific and similar to other central nervous system diseases, the diagnosis of TBE has to be established in the laboratory. The methods of choice for the diagnosis of TBE are serological assays (6). However, interpretation of serologic test results is hampered by the high cross reactivity of the antigenic structure among orthoflaviviruses, especially in areas where other orthoflaviviruses co-circulate or where vaccination against other orthoflaviviruses is regularly used (3). Examples of other medically relevant orthoflaviviruses include yellow fever virus (YFV; Orthoflavivirus flavi), dengue virus (DENV; Orthoflavivirus denguei), Zika virus (ZIKV; Orthoflavivirus zikaense), West Nile virus (WNV; Orthoflavivirus nilense), and Japanese encephalitis virus (JEV; Orthoflavivirus japonicum). In Europe, WNV is endemic (7), whereas YFV, JEV, DENV, and ZIKV infections are mainly described in association with travels to endemic areas (7–10). However locally acquired cases of DENV have been described in recent years in different European countries, including France in 2022 (11) and in Italy in 2023 (12). Vaccines against JEV and YFV are administered prior to travelling to endemic areas, and simultaneous administration of multiple vaccines is not uncommon (13, 14).

The antibody response to TBE is primarily targeted against the E and NS1 proteins of TBEV; most neutralizing antibodies recognize the viral E protein (1, 15). In order to differentiate neutralizing antibodies from cross-reactive non-neutralizing antibodies, plaque reduction neutralization, or microneutralization tests are typically used for medically relevant orthoflaviviruses, including TBEV. However, these tests are labor-intensive, time consuming, and involve the handling of infectious virus.

Reporter virus particles (RVPs) have been used in neutralization assays to measure antibodies against several orthoflaviviruses including DENV, YFV, JEV, and WNV (16–25). In particular, a system using RVPs encapsidating a sub-genomic replicon capable of expressing a reporter gene following infection of target cells has been described. In this approach, production of RVPs is accomplished by complementation of replicon RNA with orthoflavivirus structural genes expressed in trans (26). Here, we present a comprehensive assessment of the TBEV RVP seroneutralization test (SNT) using clinical samples in direct comparison with ELISA and virus-based SNT. In addition, we simultaneously assessed the neutralization capacity of sera against YFV, DENV-2, ZIKV, WNV, and JEV using RVP-based SNT assays. Our results underscore the shortcomings of traditional ELISA testing, which sometimes yielded positive results for sera incapable of neutralizing TBEV. In contrast, RVP-based SNT proved to be a robust alternative, comparable to virus-based SNT, and it can be performed under BSL2 conditions. Notably, this method offers a significant advantage of delivering results within 48h. Furthermore, cross-testing of serum samples against other orthoflaviviruses provides a valuable means to directly discern the specificity of antibodies within a given serum sample. These results argue for clinical implementation of RVP-based seroneutralization testing of TBEV and other orthoflaviviruses.

## METHODS

### Cell lines

293T, 293F, A549, Vero, BHK21 and HUH 7 cells were obtained from the laboratory of Sylvia Rothenberger (University of Lausanne, Lausanne) and Caroline Tapparel (University of Geneva, Geneva) and cryopreserved at −196°C in liquid nitrogen. Individual aliquots were thawed and serially passaged for a maximum of 15 passages. All cell lines were maintained in DMEM (1X) + GlutaMAX^TM^ + 4.5 g/L D-Glucose + 110 mg/L Sodium Pyruvate (Gibco, ThermoFisher Scientific) supplemented with 10% of fetal bovine serum (FBS, Pan Biotech) and penicillin/streptomycin 100 UI/ml (Gibco, ThermoFisher Scientific), in 75 m^2^ culture flasks (Sigma Aldrich) at 37°C in an incubator with 5% CO_2_. Cell suspensions were prepared by removing the cell culture medium from confluent flasks and detaching the cells with 0.05% Trypsin EDTA (Gibco, ThermoFisher Scientific). Cells were counted using trypan blue staining (Sigma Aldrich) and a Neubauer counting chamber.

### Viruses

TBEV strain Neudoerfl, strain Hypr, and YFV 17D, were kindly provided by Olivier Engler, Spiez Laboratory, Switzerland. TBEV was grown on A549 cells, YFV on Vero cells. Viruses were tittered by plaque assay on the cell line used for viral production, by infecting cells preplated in 24 well plates at a density of 90,000 cells/well, with serial dilutions of viruses for 1h at 37°C, followed by removal of the inoculum and addition of 500 μl of DMEM supplemented with methylcellulose 0.5%. The cells were fixed with formaldehyde 4% (Sigma Aldrich) at 72hpi and stained with crystal violet (Sigma Aldrich). Viral stocks were stored at - 80°C.

### Plasmids

The plasmids containing the CprME sequence of TBEV Neudoerfl (27), ZIKV H/PF/2013 (28), DENV-2 (29) were kindly provided by the Rockefeller University. The DENV2, WNV CprME and the pWNVII-Rep-Ren-IB replicon encoding the Renilla luciferase gene were generously given by Theodore C. Pierson from the National Institute of Health (NIH, USA) (26). The CprME plasmid of JEV (17) was kindly provided by Steve Whitehead, NIH, USA. Plasmids were propagated using MAX Efficiency™ Stbl2™ Competent Cells (ThermoFisher Scientific) according to manufacturer instructions. Extraction and purification of plasmids from transformed bacteria was performed using the PureLink™ HiPure Plasmid Midiprep Kit (ThermoFisher Scientific) and following manufacturer’s instructions.

The YFV (strain Asibi) CprME expression construct was generated as follows: The CMV promoter region from pZIKV/HPF/CprME (plasmid described in (30), and obtained from Ted Pierson, NIH) was amplified by PCR using primers RU-O-24611 and RU-O-24677. The region encoding Asibi CprME was amplified from the plasmid pACNR-2015FLYF-Asibi (PMC7426275, and GenBank MT093734) using oligos RU-O-24676 and RU-O-24678. After gel purification of the PCR products, they were joined by assembly PCR and amplified using oligos RU-O-24611 and RU-O-24678. The resultant PCR product was gel purified, digested with SnaBI and SacII, and ligated overnight at 16°C using Toyobo Ligation High V2 ligase into similarly digested and alkaline phosphatase treated pZIKV/HPF/CprME to generate pYFV/Asibi/CprME. The ligation reaction was transformed into MC1061 E. coli cells, and colonies selected on LB/carbenicillin plates for 20 h at 30°C. The sequence of the construct was verified by Sanger sequencing using oligos RU-O-19736, RU-O-18162, RU-O-23860, RU-O-24105, and RU-O-22695.

**Table 1.**
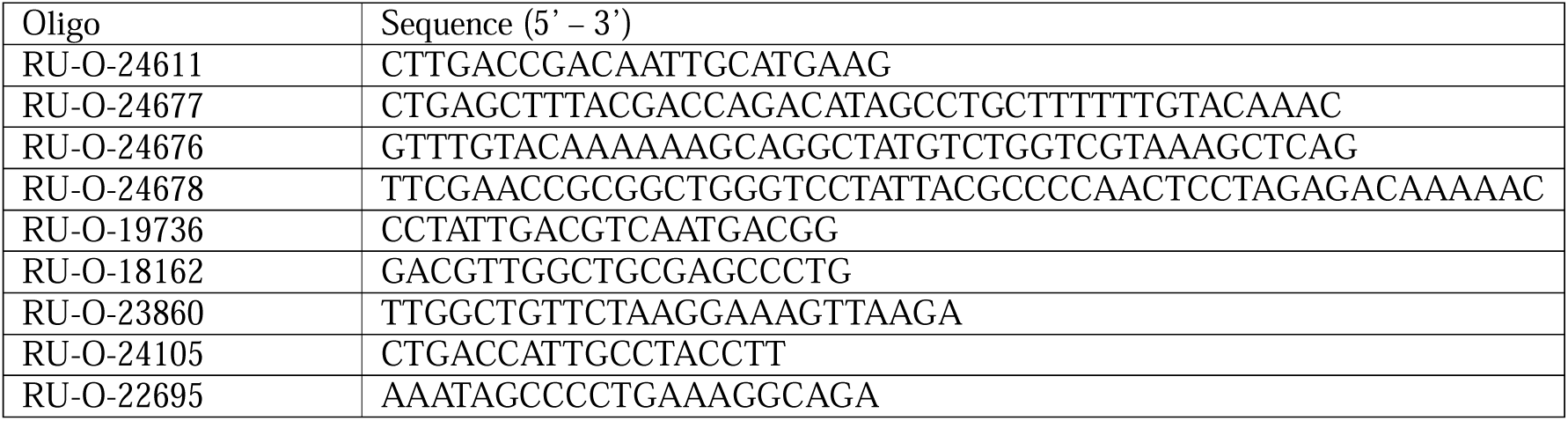
Oligos for YFV CprME cloning.

### RVP production and titration

Co-transfection of the sub-genomic replicon WNVII-Rep-Ren-IB with a plasmid encoding orthoflavivirus structural genes allows its replication, expression of luciferase, and packaging by the structural proteins provided in trans to generate RVPs. These RVPs, expressing the structural proteins of a specific orthoflavivirus (TBEV, ZIKV, DENV, WNV, JEV), can be used for a single round of infection. RVPs were produced in 293T cells, seeded the day before the transfection at 1×10^6 cells/well in poly-D-lysine-coated (ThermoFisher Scientific) 6-well plates. Three μg of CprME plasmid and 1μg of the sub-genomic replicon pWNVII-Rep-Ren- IB were co-transfected using lipofectamine 3000 (ThermoFisher Scientific) following manufacturer’s protocol. Lipid-DNA complexes were removed after 4h incubation at 37°C and replaced with cell culture medium. The culture medium containing RVPs was collected after 72-120 hours of incubation at 30°C, filtered through 0.2 μm filters (Sigma, Aldrich), and stored at −80°C in single-use aliquots.

Titration of RVPs was done by infecting HuH-7 cells plated in 96 well plates at a density of 1×10^4 cells/well with serial dilutions of RVPs at 37°C and assessing the luminescence 48 hours post infection (hpi). Luminescence was revealed with the Renilla-Glo® Luciferase Assay System (Promega) following manufacturer’s protocol. The luciferase signal was assessed with a luminometer (TriStar LB 941 by Berthold Technologies and the MikroWin 2000 software).

### Serum samples

Fifty-three clinical serum samples were provided by the University Hospital of Lausanne, Switzerland. They had been sent to the diagnostic labs for routine testing for TBEV IgG and IgM antibodies. The sera were heat-inactivated at 56°C for 30 min and were stored at −80°C. Additional sera (n=41) negative or positive for IgG and/or IgM antibodies for different orthoflaviviruses were obtained from the diagnostic laboratory ADMED Microbiologie, Switzerland, the University Hospital of Geneva, Switzerland, a ring trial organization (INSTAND, Germany) and Seracare (LGC clinical diagnostic). These serum samples were used for an initial evaluation of the RVP SNT assay.

### TBEV IgG and IgM ELISA

TBEV-specific IgG and IgM antibodies were quantified using the VIROTECH TBE IgG/IgM ELISA kit (Virotech Diagnostic’s GmbH, Germany) according to the manufacturer’s instructions. OD measurements at 450/620 nm were done using the Biotek 800/TS plate reader, the calculations of the Virotech Units (VU) were then done manually according to the manufacturer’s instructions. The cutoff for borderline results was fixed at >9 VU and the cutoff for positive results at >11 VU.

### Selection of suitable cell lines

With the goal of identifying a common cell line for both RVP- and virus-based TBEV SNT, several cell lines were tested for their ability to support infection by RVP and infectious virus. The RVP infection was assessed by infecting cells with equal amounts of RVPs and evaluating the luciferase expression at 48 hpi. The authentic virus infection was assessed by monitoring the cytopathic effect of TBEV using cell viability assays. For this, 293T and HuH- 7 cells were either mock-infected or infected with the wild type (WT) virus. 293T cells were seeded at 2×10^4 cells/well while HuH-7 cells at 1×10^4 cells/well in 96 well plates. Cells were infected with 100 plaque-forming units (PFU) of TBEV Hypr or Neudoerfl. At 72 hpi, 96 hpi and 120 hpi, the culture medium was discarded, and the cells were washed with DMEM (1X) without serum. Then, 50 μL of MTT (Sigma Aldrich) diluted at 1:10 in DMEM (1X) without serum was applied in each well. Cells were incubated at 37°C for 2h. The supernatant was then discarded and replaced with 100 μL of dimethyl sulfoxide (DMSO; Sigma Aldrich) per well to lyse the cells. Absorbance of cells was measured at 570 nm with a spectrophotometer.

### RVP serum neutralization test

In order to facilitate simultaneous evaluation of serum samples for their neutralizing capacity against various orthoflaviviruses, we established an RVP SNT protocol that can be applied uniformly to all RVPs used in this study (TBEV, YFV, ZIKV, DENV, WNV, JEV). The day before the assay, HuH-7 cells were seeded in a 96-well plate at a density of 1×10^4 cells/well. The RVPs were diluted in DMEM supplemented with 2.5% FBS to a concentration of 1×10^4 RLU/well. 60 µL of RVPs and 60 µL of serum samples serially diluted 4-fold in DMEM supplemented with 2.5% FBS were mixed in a 96 well V bottom plate and incubated for 1h at 37°C. Thereafter, the medium was removed from the cells in the 96-well plates, and the RVP / serum sample mixture was added to the cells. A line of untreated control (containing RVP and medium) and a line of uninfected control (containing only cell medium) were included in every plate. This plate was then incubated for 48h at 37°C. To determine the results of neutralization assays, 70 μL of supernatant was discarded from each well and replaced with 50 μL of Renilla-Glo® Luciferase Assay System (Promega). The luciferase signal was assessed with a luminometer (TriStar LB 941 by Berthold Technologies and the MikroWin 2000 software). The concentration at which 99% of cell infection by the RPVs was inhibited (IC_99_) was determined with nonlinear regression with GraphPad by calculating the percentages of infection at different dilutions compared to the infected untreated (i.e., no serum sample added) control.

### Wild type virus serum neutralization test

As a reference method for TBEV, we used a standard WT virus serum neutralization test (virus-based SNT) to determine the neutralizing activity of serum samples. Similarly, results obtained using the YFV RVP SNT were compared to those obtained using a YFV WT SNT. All work with infectious viruses was performed in a BSL-3 laboratory. 96-well plates were seeded with 1×10^4 HuH-7 cells/well the day before infection. 60 µl of serum samples serially diluted 4-fold were mixed with 60 µL of WT virus suspension, corresponding to 100 PFU/well. After an incubation period of 1h at 37°C, the serum sample / virus mixture was added to the cells in the 96-well plates, from which the medium had been removed. As for the RVP neutralization assay, a line of untreated control (containing WT virus and medium) and a line of uninfected control (containing only cell medium) were included in every plate. The plate was incubated at 37° for 4 (TBEV Hypr, YFV) to 5 days (TBEV Neudoerfl). Cytopathic effect was assessed by coloration of cells with crystal violet and optical microscopy. The IC99 was determined based on the last dilution in which the sera showed complete protection from cytopathic effect. The cutoff for a positive WT SNT result was set at >1:8.

### RVP SNT assay characteristics and cutoff definition

The sensitivity and specificity of the TBEV RVP SNT were defined based on receiver operating curve (ROC) analysis, for which we used the TBEV WT SNT results as a reference standard. The cutoff for qualitative test evaluation (positive/negative) was defined based on the maximal Youden’s index (31). This cutoff value was implemented consistently across all RVP SNTs.

### Correlation analysis

We utilized a linear regression model to assess the correlation between the quantitative RVP SNT and WT SNT results, as well as between the RVP SNT and the ELISA as well as the WT SNT and the ELISA results. Graphs were constructed by reporting the log of the 1/IC99 of each serum for the RVP SNT, the WT SNT, or the IgG or IgM ELISA. All analyses were performed using GraphPad Prism 9, a p value < 0.05 was considered as significant.

### Ethics approval

Ethical authorization to re-use samples collected by the serology of CHUV has been granted by CER-VD, project n°2023-01971.

### Data sharing

Raw data of SNT are available upon reasonable requests to

## RESULTS

### Optimization of SNT conditions

With the goal to identify a common cell line for both RVP-based and virus-based SNT for TBEV, three cell lines were tested for their ability to express the reporter gene following RVP infection (Supplementary Figure 1A) and their potential to show a cytopathic effect upon WT virus infection (Supplementary Figure 1B-C). TBEV stocks were produced in A549 cells. However, these cells yielded poor luminescence signal following RVP infection compared to other cell lines, indicating limited expression of replicon in these cells (Supplementary Figure 1A). Among the permissive cell lines, HuH-7 showed the best profile in both assays and was therefore used in all subsequent experiments. These cells were also susceptible to RVPs derived from WNV, JEV, YFV, DENV, and ZIKV (Supplementary Figure 1D).

Next, we evaluated the performance of RVP-based SNT assays using three to seven serum samples for each of the TBEV, WNV, JEV, YFV, DENV, and ZIKV and two samples negative for antibodies against the respective orthoflaviviruses included in this study (TBEV, WNV, JEV, YFV, DENV, and ZIKV). For all positive samples, we were able to calculate the IC_99_ values using the respective RVP SNT assay, whereas none of the negative samples exhibited neutralizing activity against their respective RVPs (Supplementary Figure 2).

### RVP SNT assay characteristics and cutoff definition

The cutoff for qualitative test evaluation (negative/positive) of the TBEV RVP-based SNT was defined by ROC analysis using TBEV virus-based SNT results as a reference. Youden’s index was maximal at a value of 1:40, with a test sensitivity of 91.7% (95% CI: 87.2-97.1%) and a test specificity of 100% (95% CI: 79.6-100%) (Supplementary Figure 3). The cutoff value of ≥1:40, defined using this approach, was implemented consistently across all RVP SNTs.

### RVP-based SNT and virus-based SNT results for TBEV

A total of 53 serum samples for which TBEV-specific IgG and IgM titers had been assessed by ELISA-based routine testing (Virotech TBE IgG/IgM ELISA kit) were analyzed using RVP-based and virus-based (strain Neudoerfl) SNT assays. The results are shown in Table 2. 94.3% of samples (50/53) yielded concordant results with both RVP-based and virus-based SNT assays; the remaining 3 samples with discordant results are probably false-negative in the RVP-based SNT as compared to TBEV WT SNT. The results of the RVP-based assay differed from those of ELISA for 12 samples (22.6%). Of these, two samples were positive by RVP-based assay but negative or borderline by ELISA; both samples were confirmed positive by the virus-based SNT assay. Ten samples tested negative in RVP-based assay but positive in TBEV IgG ELISA (7 negative, 1 borderline, and 2 positive for IgM), whereof 8 were confirmed negative by virus-based SNT.

**Table 2.** Anti-TBEV antibody test results of clinical samples. ^1^ Virotech TBE IgG/IgM ELISA; results are given in Virotech Units / ml; cutoffs for negative and positive results were set at ≤9 and ≥11 VU/ml, respectively. Values not divisible by 4 are obtained by the mean of two independent SNT; ^2^ TBEV reporter virus particle seroneutralization test; results are given as IC99; the cutoff for a positive result was set at ≥1:40; ^3^ TBEV wild type seroneutralization test; results are given as IC99; the cutoff for a positive result was set at >1:8; ^4^ discordance of qualitative results between different tests are indicated with x. For this comparison, ELISA was regarded as positive if either IgG or IgM or both tested positive; borderline ELISA results were regarded as negative.

|  | ELISA<sup>1</sup> |  |  |  | RVP SNT<sup>2</sup> |  | WT SNT<sup>3</sup> |  | discording qualitative results<sup>4</sup> |  |  |
| --- | --- | --- | --- | --- | --- | --- | --- | --- | --- | --- | --- |
| sample | IgG<br>qn | IgG<br>ql | IgM<br>qn | IgM<br>ql | Ig tot.<br>qn | Ig tot.<br>ql | Ig tot.<br>qn | Ig tot.<br>ql | RVP SNT<br>/ WT SNT | RVP SNT<br>/ ELISA | WT SNT<br>/ ELISA |
| CH01 | <9 | n | <9 | n | <1:8 | n | <1:8 | n |  |  |  |
| CH02 | <9 | n | <9 | n | <1:8 | n | <1:8 | n |  |  |  |
| CH03 | <9 | n | <9 | n | <1:8 | n | <1:8 | n |  |  |  |
| CH04 | <9 | n | <9 | n | <1:8 | n | <1:8 | n |  |  |  |
| CH05 | <9 | n | <9 | n | <1:8 | n | <1:8 | n |  |  |  |
| CH06 | <9 | n | <9 | n | <1:8 | n | <1:8 | n |  |  |  |
| CH07 | <9 | n | <9 | n | <1:8 | n | <1:8 | n |  |  |  |
| CH08 | <9 | n | <9 | n | <1:8 | n | <1:8 | n |  |  |  |
| CH09 | <9 | n | <9 | n | 1:10 | n | 1:32 | p | x |  | x |
| CH10 | <9 | n | <9 | n | 1:96 | p | 1:32 | p |  | x | x |
| CH11 | <9 | n | <9 | n | 1:45 | p | 1:32 | p |  | x | x |
| CH12 | <9 | n | 14 | p | 1:49 | p | 1:128 | p |  |  |  |
| CH13 | 10 | ? | <9 | n | 1:39 | n | 1:8 | n |  |  |  |
| CH14 | 17 | p | <9 | n | <1:8 | n | <1:8 | n |  | x | x |
| CH15 | 21 | p | <9 | n | 1:12 | n | 1:8 | n |  | x | x |
| CH16 | 32 | p | <9 | n | <1:8 | n | 1:8 | n |  | x | x |
| CH17 | 35 | p | <9 | n | <1:8 | n | <1:8 | n |  | x | x |
| CH18 | 24 | p | <9 | n | 1:17 | n | 1:8 | n |  | x | x |
| CH19 | 51 | p | <9 | n | <1:8 | n | 1:8 | n |  | x | x |
| CH20 | 40 | p | <9 | n | <1:8 | n | <1:8 | n |  | x | x |
| CH21 | 24 | p | <9 | n | 1:398 | p | 1:80 | p |  |  |  |
| CH22 | 32 | p | <9 | n | 1:57 | p | 1:128 | p |  |  |  |
| CH23 | 60 | p | <9 | n | 1:336 | p | 1:104 | p |  |  |  |
| CH24 | 34 | p | <9 | n | 1:105 | p | 1:32 | p |  |  |  |
| CH25 | 60 | p | <9 | n | 1:559 | p | 1:320 | p |  |  |  |
| CH26 | 45 | p | <9 | n | 1:588 | p | 1:2720 | p |  |  |  |
| CH27 | 26 | p | <9 | n | 1:533 | p | 1:200 | p |  |  |  |
| CH28 | 38 | p | <9 | n | 1:55 | p | 1:80 | p |  |  |  |
| CH29 | 37 | p | <9 | n | 1:64 | p | 1:320 | p |  |  |  |
| CH30 | 44 | p | <9 | n | 1:83 | p | 1:32 | p |  |  |  |
| CH31 | 23 | p | <9 | n | 1:125 | p | 1:32 | p |  |  |  |
| CH32 | 12 | p | <9 | n | 1:85 | p | 1:32 | p |  |  |  |
| CH33 | 12 | p | <9 | n | 1:54 | p | 1:32 | p |  |  |  |
| CH34 | 19 | p | <9 | n | 1:336 | p | 1:128 | p |  |  |  |
| CH35 | 29 | p | <9 | n | 1:47 | p | 1:128 | p |  |  |  |
| CH36 | 41 | p | <9 | n | 1:74 | p | 1:128 | p |  |  |  |
| CH37 | 25 | p | 10 | ? | <1:8 | n | 1:80 | p | x | x |  |
| CH38 | 54 | p | 10 | ? | 1:3145 | p | 1:3200 | p |  |  |  |
| CH39 | 14 | p | 14 | p | 1:16 | n | 1:8 | n |  | x | x |
| CH40 | 41 | p | 21 | p | 1:8 | n | 1:32 | p | x | x |  |
| CH41 | 32 | p | 62 | p | 1:8183 | p | 1:3200 | p |  |  |  |
| CH42 | 17 | p | 13 | p | 1:215 | p | 1:200 | p |  |  |  |
| CH43 | 38 | p | 100 | p | 1:3921 | p | 1:2600 | p |  |  |  |
| CH44 | 17 | p | 45 | p | 1:1971 | p | 1:1280 | p |  |  |  |
| CH45 | 24 | p | 51 | p | 1:1641 | p | 1:2048 | p |  |  |  |
| CH46 | 34 | p | 21 | p | 1:40 | p | 1:512 | p |  |  |  |
| CH47 | 49 | p | 22 | p | 1:364 | p | 1:512 | p |  |  |  |
| CH48 | 30 | p | 65 | p | 1:2002 | p | 1:3584 | p |  |  |  |
| CH49 | 36 | p | 67 | p | 1:1579 | p | 1:512 | p |  |  |  |
| CH50 | 34 | p | 45 | p | 1:833 | p | 1:896 | p |  |  |  |
| CH51 | 47 | p | 50 | p | 1:143 | p | 1:896 | p |  |  |  |
| CH52 | 15 | p | 50 | p | 1:7032 | p | 1:12800 | p |  |  |  |
| CH53 | 38 | p | 31 | p | 1:291 | p | 1:128 | p |  |  |  |

Linear regression analysis was used to assess the agreement between quantitative test results obtained from RVP-based versus virus-based (strain Neudoerfl) SNT, as well as between SNTs and ELISA (Figure 1). All correlations were significant (p<0.001, T test for linear regression). A high correlation coefficient (R^2^ 0.86) was obtained for RVP-based SNT versus virus-based SNT (Figure 1A), RVP-based SNT versus IgM ELISA (R^2^ 0.89), and virus-based SNT versus IgM ELISA (R^2^ 0.91) (Figure 1D, 1E).

**Figure 1.**
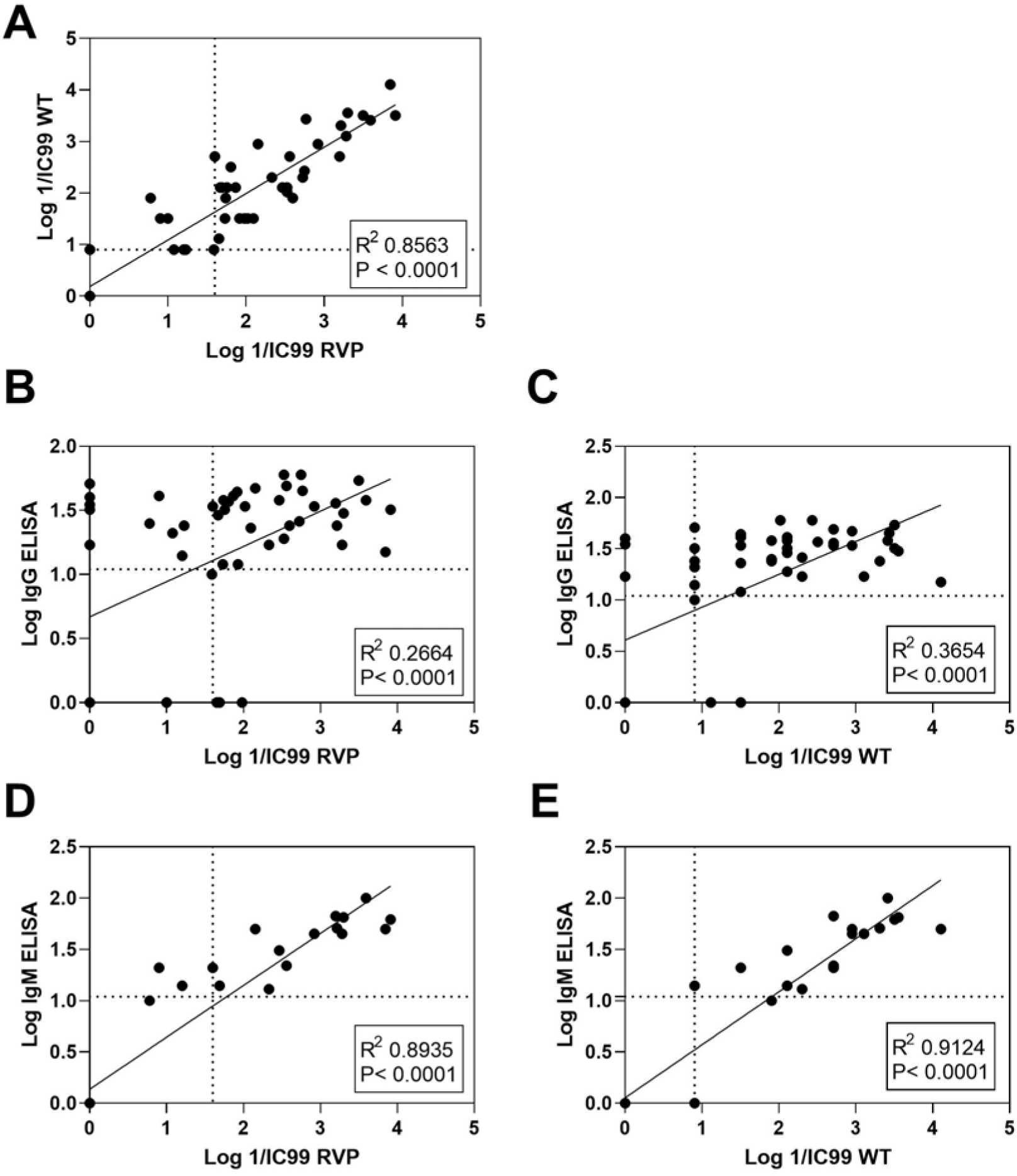
Linear regression analysis comparing results obtained using the TBEV RVP SNT, the TBEV WT SNT, and TBEV IgG and IgM ELISA. SNT results are expressed as log 1/IC99, ELISA results as log VU/ml. A) correlation between the TBEV RVP SNT and the TBEV WT SNT (strain Neudoerfl). B) correlation between the TBEV RVP SNT and TBEV IgG ELISA results. C) correlation between the TBEV WT SNT and TBEV IgG ELISA results. D) correlation between the TBEV RVP SNT and TBEV IgM ELISA results. E) correlation between TBEV WT SNT and TBEV IgM ELISA results. Linear regression was calculated with GraphPad Prism, correlation coefficient and p values (T test for linear regression) are indicated in each panel. Dotted lines represent the cutoff of the respective tests.

### Impact of virus strain on virus-based SNT results

For a subset of samples, we evaluated the concordance between RVP-based and virus-based SNT results when using a different virus strain, Hypr, in the virus-based SNT. Qualitatively, the virus based SNT results were identical in 17 out of 18 samples (94.4%) when using the strains Neudoerfl and Hypr, while quantitative results showed some variation (Supplementary Table 1). The linear regression comparing RVP (strain Neudoerfl) and virus-based (strain Hypr) SNT results R^2^ of 0.55, as compared to the R^2^ value of 0.86 when comparing RVP-based and virus-based (strain Neudoerfl) SNT, respectively (Supplementary Figure 4).

### Evaluation of selected sera for crossreactivity

The results of the RVP-based assay differed from those of ELISA for 12 samples (22.6%). These samples, one borderline in ELISA and negative in the RVP-based assay, and 7 additional, randomly selected samples were tested for their neutralizing potential against RVPs of 5 different orthoflaviviruses (ZIKV, YFV, DENV, WNV, JEV). The results are shown in Table 3. Nine of the 10 serum samples (90%) that were TBEV-positive by ELISA but negative by RVP-based SNT tested positive for neutralizing antibodies against other orthoflaviviruses. Moreover, we found that seven of the 15 ELISA-positive samples (46.7%) contained neutralizing antibodies against multiple orthoflaviviruses.

**Table 3.** RVP SNT results against multiple orthoflaviviruses. ^1^ Virotech TBE IgG/IgM ELISA; results are given in Virotech Units / ml; cutoffs for negative and positive results were set at ≤9 and ≥11 VU/ml, respectively; ^2^ reporter virus particle seroneutralization test; results are given as IC99; the cutoff for a positive result was set at ≥1:40. n.a. not assessed

|  | ELISA<sup>1</sup> |  |  |  | TBEV<br>RVP SNT<sup>2</sup> |  | ZIKV<br>RVP SNT<sup>2</sup> |  | YFV<br>RVP SNT<sup>2</sup> |  | DENV<br>RVP SNT<sup>2</sup> |  | WNV<br>RVP SNT<sup>2</sup> |  | JEV<br>RVP SNT<sup>2</sup> |  | Clinical information |
| --- | --- | --- | --- | --- | --- | --- | --- | --- | --- | --- | --- | --- | --- | --- | --- | --- | --- |
| sample | IgG<br>qn | IgG<br>ql | IgM<br>qn | IgM<br>ql | Ig tot. qn | Ig tot. ql | Ig tot. qn | Ig tot. ql | Ig tot. qn | Ig tot. ql | Ig tot. qn | Ig tot. ql | Ig tot. qn | Ig tot. ql | Ig tot. qn | Ig tot. ql |  |
| CH41 | 32 | p | 62 | p | 1:8183 | p | < 1:8 | n | 1:13 | n | < 1:8 | n | < 1:8 | n | < 1:8 | n | Diagnosed TBE case |
| CH43 | 38 | p | 100 | p | 1:3921 | p | < 1:8 | n | 1:134 | p | 1:47 | p | < 1:8 | n | < 1:8 | n | Diagnosed TBE case |
| CH21 | 24 | p | < 9 | n | 1:398 | p | < 1:8 | n | 1:659 | p | < 1:8 | n | 1:48 | p | 1:77 | p | Vaccinated against<br>TBEV, JEV, YFV |
| CH22 | 32 | p | < 9 | n | 1:57 | p | < 1:8 | n | 1:185 | p | < 1:8 | n | 1:20 | n | 1:649 | p | Vaccinated against<br>TBEV, JEV, YFV |
| CH23 | 60 | p | < 9 | n | 1:336 | p | < 1:8 | n | 1:1132 | p | < 1:8 | n | < 1:8 | n | < 1:8 | n | Vaccinated against<br>TBEV, YFV |
| CH39 | 14 | p | 14 | p | 1:16 | n | < 1:8 | n | 1:90 | p | < 1:8 | n | 1:36 | n | < 1:8 | n | Travel in Mexico, South Africa<br>Thailand, Peru |
| CH37 | 25 | p | 10 | ? | < 1:8 | n | < 1:8 | n | 1:112 | p | < 1:8 | n | 1:145 | p | 1:29 | n | Neurological symptoms |
| CH40 | 41 | p | 21 | p | 1:8 | n | < 1:8 | n | 1:1557 | p | 1:12 | n | < 1:8 | n | < 1:8 | n | No data |
| CH14 | 17 | p | < 9 | n | < 1:8 | n | < 1:8 | n | 1:426 | p | n.a. |  | n.a. |  | n.a. |  | Origin Vietnam |
| CH15 | 21 | p | < 9 | n | 1:12 | n | < 1:8 | n | 1:1786 | p | < 1:8 | n | < 1:8 | n | < 1:8 | n | Travel in Brazil, Thailand,<br>Mexico |
| CH16 | 32 | p | < 9 | n | < 1:8 | n | < 1:8 | n | 1:87 | p | 1:17 | n | < 1:8 | n | < 1:8 | n | Meningo-encephalitis of<br>unknown etiology |
| CH18 | 24 | p | < 9 | n | 1:17 | n | < 1:8 | n | < 1:8 | n | < 1:8 | n | 1:40 | p | < 1:8 | n | Neurological symptoms<br>associated to glioblastoma |
| CH20 | 40 | p | < 9 | n | < 1:8 | n | < 1:8 | n | 1:54 | p | 1:102 | p | < 1:8 | n | 1:20 | n | Origin Indonesia |
| CH17 | 35 | p | < 9 | n | < 1:8 | n | < 1:8 | n | 1:1588 | p | 1:62 | p | 1:97 | p | < 1:8 | n | Diagnosed WNV infection.<br>Tested positive for IgG for YFV<br>DENV and WNV. Tested<br>positive for IgM for WNV. |
| CH19 | 51 | p | < 9 | n | < 1:8 | n | 1:8 | n | < 1:8 | n | < 1:8 | n | < 1:8 | n | < 1:8 | n | Meningo-Encephalitis of<br>unknown etiology |
| CH13 | 10 | ? | < 9 | n | 1:39 | n | < 1:8 | n | 1:22 | n | < 1:8 | n | < 1:8 | n | 1:11 | n | Vaccinated with two doses of<br>TBEV vaccine |
| CH10 | < 9 | n | < 9 | n | 1:96 | p | < 1:8 | n | < 1:8 | n | < 1:8 | n | < 1:8 | n | < 1:8 | n | No data |
| CH11 | <9 | n | <9 | n | 1:45 | p | < 1:8 | n | 1:377 | p | < 1:8 | n | 1:11 | n | 1:36 | n | Vaccinated with one dose of TBEV - No vaccination for YF |
| CH06 | <9 | n | <9 | n | < 1:8 | n | < 1:8 | n | < 1:8 | n | < 1:8 | n | < 1:8 | n | < 1:8 | n | No data |
| CH09 | <9 | n | <9 | n | 1:10 | n | < 1:8 | n | < 1:8 | n | < 1:8 | n | < 1:8 | n | < 1:8 | n | Vaccinated with two doses of TBEV vaccine – immunosuppressed patient |

Clinical information was available for 17/20 samples. Thereof, 12 were in accordance with (samples CH41, CH22, CH23, CH15, CH20, CH17) or partially matched the SNT results (samples CH43, CH21, CH37, CH39, CH15, CH20, CH11), whereas 5 did not provide sufficient evidence to explain the results obtained (samples CH14, CH19, CH09, CH18, CH16).

Twelve of the 15 samples (80%) that were positive by TBEV IgG ELISA showed a neutralizing activity against YFV. This percentage, although partly supported by the clinical data, is surprisingly high when compared to the assumed seroprevalence of YFV in the population. Nonetheless, the presence of YFV antibodies in these samples was confirmed by both RVP-based and virus-based SNT (Supplementary Table 2). with full qualitative concordance between the two methods.

## DISCUSSION

Despite the availability of effective vaccines, TBEV continues to pose a considerable threat to public health due to its expanding geographical distribution and the increasing number of reported cases (1). Serological diagnosis plays a crucial role in the management and surveillance of TBEV infections. However, it is fraught with challenges, primarily stemming from the cross-reactivity among orthoflaviviruses. This cross-reactivity can result in false-positive results and misdiagnosis, especially in regions where multiple orthoflaviviruses co-circulate, for persons having traveled to areas endemic for other orthoflaviviruses, or for persons recently vaccinated against other orthoflaviviruses (3). While most laboratories use ELISA for routine serological diagnosis, a reliable differentiation of virus-neutralizing from cross-reactive virus-binding but non-neutralizing antibodies is only possible using serum neutralization tests. Here, we describe the evaluation of an RVP-based test system, allowing for the simultaneous detection of antibodies directed against multiple orthoflaviviruses. The RVP-based SNT can serve as a rapid (within 48 hours) substitute for the virus-based SNT, requiring only BSL-2 facilities and demonstrating a reliable performance. The concurrent identification of antibodies against other orthoflaviviruses holds significant importance.

RVP-based SNTs for different orthoflaviviruses have been established using different cell lines, including BHK-J cells (24), BHK-21 cells (26), Vero cells (25), and HUH-7.5 cells (27). In our experiments, we found that HUH7 cells were well suited for the effective execution of TBEV, YFV, DENV, ZIKV, WNV, and JEV RVP SNTs. The readout of our assay using Renilla luciferase was performed 48hpi, which is in line with the readout of a test using Gaussia luciferase (32), while others described readout times of 3 to 5-6 days using NanoLuc or GFP, respectively(33, 34).

A major difficulty when establishing diagnostic tests is the definition of an appropriate cutoff, which discriminates positive from negative results and requires a compromise between sensitivity and specificity. We performed an ROC analysis and set the cutoff point for the TBEV RVP-based SNT at the value resulting in the highest Youden’s index, maximizing the summation of sensitivity and specificity (31). One of the possible limitations of our study is that cutoff values were not defined for each orthoflavivirus RVP SNTs individually, but rather by applying the cutoff defined for the TBEV RVP SNT to all other test systems While this offers the opportunity of harmonizing the readout of all tests, sensitivity and / or specificity of RVP-based SNTs other than TBEV might not be optimal. Due to the small sample size, an individual cutoff definition based on ROC analysis for each individual RVP SNT was not possible in this study.

When compared to the virus-based SNT (strain Neudoerfl), the RVP-based SNT showed a sensitivity of 91.7% and a specificity of 100% for TBEV. Assay specificity is of high importance for a diagnostic assay that may be used as a confirmatory test. An assay sensitivity of 91.7% is acceptable, though not optimal. On the other hand, we did also observe positive results by RVP-based SNT for samples testing negative in ELISA (samples CH10, CH11, Table 2), which were confirmed positive by the virus-based SNT. Thus, while ELISA tests for the detection of anti-TBEV IgG antibodies generally show a high sensitivity, reaching up to 99% (35), it is important to acknowledge the potential for false-negative outcomes in these assays. Given the quick availability of results with the RVP-based SNT, opting for this test may offer advantages.

Discording results were obtained for 22.6% of samples (12/53) when comparing the RVP-based SNT to ELISA. Thereof, 10 were false-positive in ELISA. The pronounced cross-reactivity of antibodies against different orthoflaviviruses in ELISA test systems is well known (3). In fact, we could demonstrate the presence of antibodies against other orthoflaviviruses in 9/10 samples yielding false-positive results by ELISA (Table 3). Neurological symptoms of TBE are similar to those of other viral infections of the brain, including for instance WNV, which is endemic in Europe (7). Conducting a test that can both rule out cross-reactivity and simultaneously evaluate specific seroneutralization for multiple orthoflaviviruses is thus highly clinically significant. This capability is one of the significant advantages provided by the here-described RVP-based SNTs.

Interestingly, the results of IgM ELISA showed a higher level of correlation with those from RVP-based SNT (R^2^ 0.89, Figure 1D) and virus-based SNT (R^2^ 0.91, Figure 1E) compared to those from IgG ELISA (R^2^ 0.27 and 0.37, respectively, Figure 1B and 1C). This discrepancy likely arises from the fact that a substantial number of samples that test positive solely for IgG are from individuals who have been vaccinated against different (crossreactive) orthoflaviviruses or have recovered from an earlier infection, whereas those that also test positive for IgM are typically associated with clinical TBE cases. It is also in line with the observation that tests for anti-TBEV IgM are usually more specific than IgG tests with regard to cross-reactivity with other orthoflaviviruses (36, 37).

While there was a strong correlation between RVP-based and virus-based SNT assays when the strain Neudoerfl was used in both assays (R^2^ 0.86, Figure 1A), TBEV RVP SNT results correlated less well to TBEV WT SNT results using the strain Hypr (R^2^ 0.55, Supplementary Figure 4), as expected since the TBEV RVP utilized in this study is derived from the genetic sequence of the Neudoerfl strain of TBEV. The results are in line with literature, where it has been shown that antibodies generated upon vaccination with specific strains do not equally efficiently neutralize different wild type virus strains (38, 39).

Among the subset of samples tested for their neutralizing capacity against multiple orthoflaviviruses, we found a high proportion of samples (8/20) positive for antibodies against more than one virus (Table 3). Thus, multiple vaccinations or exposures were common in this set of samples, as verified in some cases by concordance between clinical information and SNT results. However, for some sera the clinical information did not provide sufficient evidence to explain the SNT results or only partially matched the SNT results, possibly linked to an incomplete dataset or anamnesis may not have included questions about different orthoflavivirus vaccinations. Our findings underscore the importance of simultaneous testing against a large panel of orthoflaviviruses. Only by doing so can we unveil the complete range of neutralizing antibodies present in a sample.

Remarkably, a significant number of samples yielded positive results for neutralizing antibodies against YFV (Table 3). To address concerns about the specificity of the YFV RVP-based SNT, all results were verified using the virus-based SNT (Supplementary Table 2). The observed positivity rate is likely influenced by our choice to conduct RVP SNTs for multiple orthoflaviviruses on selected samples. This selection primarily relied on discordant outcomes between TBEV ELISA and TBEV RVP SNT results. Nevertheless, YFV vaccination is a non-negligible cause of false-positive TBEV (IgG) ELISA results, given the (travel) vaccination recommendations. However, it is important to consider that YFV vaccination can significantly contribute to false-positive TBEV (IgG) ELISA results, especially in light of travel vaccination guidelines (40) and the phylogenetic similarity of YFV to TBEV (12).

We found two samples positive in TBEV ELISA testing but negative in all orthoflavivirus RVP-based SNTs (TBEV, ZIKV, YFV, DENV, WNV, JEV). While this might be due to false-negative results in RVP-based SNTs, we must also keep in mind other less-well characterized orthoflaviviruses which might be of medical importance but have not been included in our study. These could be tick-borne (e.g., Powassan virus [*Orthoflavivirus powassense*], Omsk hemorrhagic fever virus [*Orthoflavivirus omskense*] or mosquito borne pathogens (e.g., Usutuvirus [*Orthoflavivirus usutuense*], Wesselsbron virus [*Orthoflavivirus wesselsbronense*]) (41, 42).

Taken together, our results emphasize the importance of performing neutralization instead of ELISA testing for diagnosing orthoflaviviral infections. While this has been labor-intensive and requiring high biosafety level standards, the RVP-based neutralization approach for multiple orthoflaviviruses we described provides a notable advantage, offering a more accessible and cost-effective platform and thus enhancing the diagnostic capabilities of healthcare institutions.

## Data Availability

All data produced in the present study are available upon reasonable request to the authors

## AKNOWLEDGEMENTS

The authors would like to thank Emilie Ruegger and Alexandre Mamin for their support in providing the sera, Adriana Renzoni for providing reference sera from the Swiss Center for Emerging viruses and the members of the Swiss National Reference Center for Tick Transmitted Diseases for fruitful discussions.

