## Supplementary data for "A reporter virus particle seroneutralization assay for tick-borne encephalitis virus overcomes ELISA limitations"

Valeria Cagno

Institute of Microbiology of Lausanne

Rue du Bugnon 48

1011 Lausanne

+41213142611

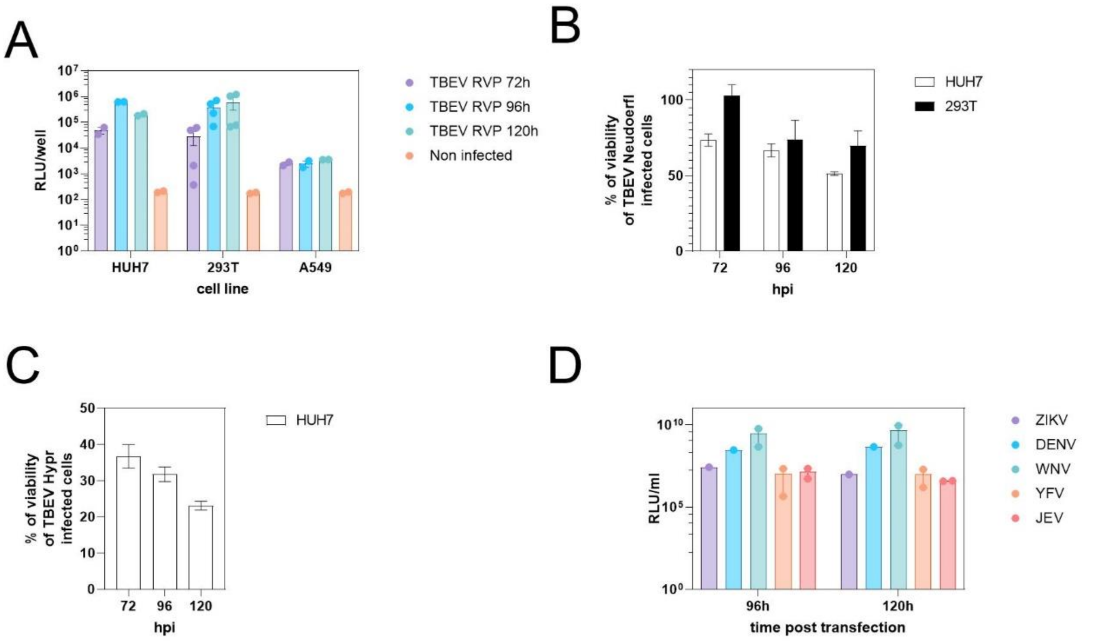


**Supplementary Figure 1. Selection of suitable cell lines.** A) suitability of HUH7, 293T, and A549 cell lines for the propagation of TBEV RVPs; the three cell lines were infected with 20 µl of TBEV RVP and, after 48 hours of incubation, luminescence was measured. The RVP collected at different time post-transfection of the 293T cells (72, 96 and 120h) were tested in parallel; B) extent of cytopathic effect produced TBEV WT strain Neudoerfl on HUH7 and 293T cells; cells were infected with 100 PFU/well and their viability assessed by a viability (MTT) assay 72, 96 or 120 hpi. C) extent of cytopathic effect produced by TBEV WT strain Hypr; the viability assay was performed as described for TBEV WT strain Neudoerfl; D) suitability of HUH7 cell lines for replication of WNV replicon with structural proteins of ZIKV, DENV, WNV, YFV, and JEV. expressed in trans; RVP collected after 96 or 120h from transfection at 30°C, were tittered on HUH7 cells and the luminescence was measured after 48h of incubation. Titers are expressed in Relative Luminescence Unit/ml

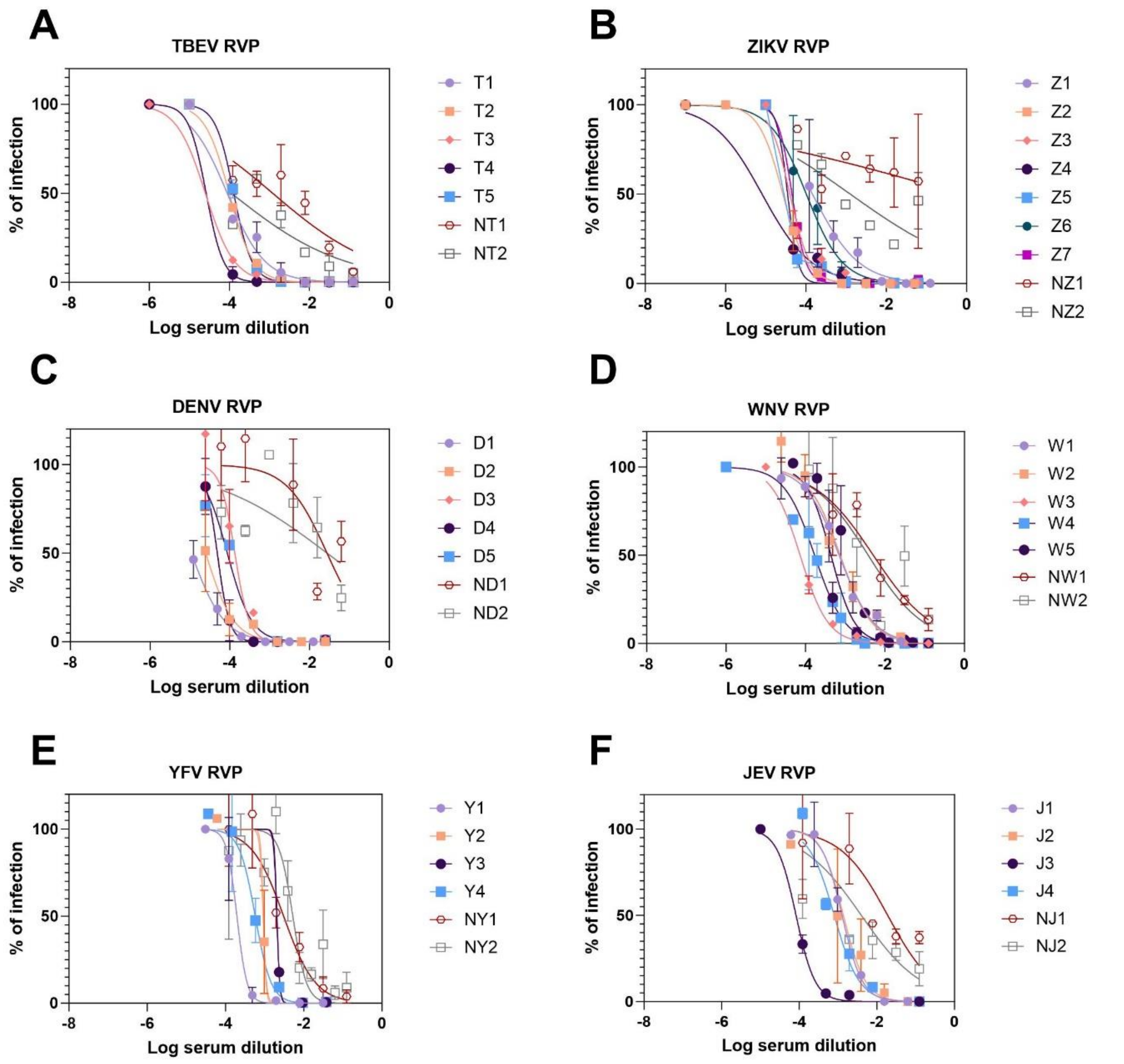


**Supplementary Figure 2. Evaluation of different RVP SNTs.** A) Evaluation of TBEV RVP SNT with TBEV IgG positive sera from commercial quality controls (T1 to T5) and sera from routine diagnostics tested negative by TBEV IgG and IgM ELISA (NT1 and NT2) B) Evaluation of ZIKV RVP SNT with ZIKV IgG and IgM positive sera from the Swiss reference center for emerging diseases (Z1 and Z2) or from commercial quality controls (Z3 to Z5), sera ZIKV IgG positive from commercial quality controls (Z6 and T7) and sera from quality controls tested negative by Zika IgG and IgM ELISA (NZ1 and NZ2) C) Evaluation of DENV RVP SNT with DENV IgG positive sera from the Swiss reference center for emerging diseases (D1 and D2) or from quality controls (D3) and IgG and IgM positive sera from commercial quality controls (D4-D5) and sera from quality controls tested negative by DENV IgG and IgM ELISA (ND1 and ND2) D) Evaluation of WNV RVP SNT with WNV IgG positive sera (W1 and W2, W3) and IgG and IgM positive sera (W4, W5) from the Swiss reference center for emerging diseases or from commercial controls and sera from routine diagnostics tested negative by WNV IgG and IgM ELISA (NW1 and NW2) E) Evaluation of YFV RVP SNT with sera of YFV vaccinated individuals (Y1 to Y4) and sera tested negative by IF for YFV (NY1 and NY2) E) Evaluation if JEV RVP SNT with sera of JEV vaccinated individuals (J1 to J4) and sera tested negative by IF for YFV (NY1 and NY2).

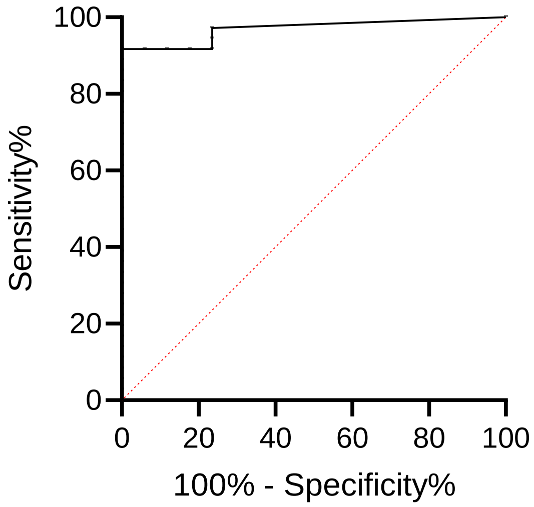


**Supplementary Figure 3. Receiver operating characteristics (ROC) analysis and calculation of Youden's index.** Each point of the black line on a ROC curve corresponds to a cut-off value and is associated with a test sensitivity and specificity. The red dotted line represents positive/negative results that are given by chance. Thus, locating the cut-off point requires a compromise between sensitivity and specificity. In this approach, we set the cutoff point at the value resulting in a maximum Youden’s index (i.e. maximum distance by the red-dotted line). The maximum Youden’s index (190.7) was reached at a titer of 1:40, with a test sensitivity of 91.7% (95% CI: 87.2-97.1%) and a test specificity of 100% (95% CI: 79.6-100%).

| | RVP SNT[1] | | WT SNT Neudoerfl[2] | | WT SNT Hypr[2] | | discording qualitative results[3] | |
|---|---|---|---|---|---|---|---|---|
| **sample** | **Ig tot. qn** | **Ig tot. ql** | **Ig tot. qn** | **Ig tot. ql** | **Ig tot. qn** | **Ig tot. ql** | **RVP SNT / WT SNT Neudoerfl** | **RVP SNT / WT SNT Hypr** |
| **CH03** | <1:8 | n | <1:8 | n | <1:8 | n | | |
| **CH04** | <1:8 | n | <1:8 | n | <1:8 | n | | |
| **CH14** | <1:8 | n | <1:8 | n | <1:8 | n | | |
| **CH15** | <1:8 | n | <1:8 | n | <1:8 | n | | |
| **CH13** | 1:39 | n | 1:8 | n | 1:8 | n | | |
| **CH28** | 1:55 | p | 1:80 | p | <1:8 | n | | x |
| **CH25** | 1:559 | p | 1:271 | p | 1:680 | p | | |
| **CH38** | 1:3145 | p | 1:3200 | p | 1:1280 | p | | |
| **CH26** | 1:588 | p | 1:2720 | p | 1:80 | p | | |
| **CH41** | 1:8183 | p | 1:3200 | p | 1:5120 | p | | |
| **CH29** | 1:64 | p | 1:320 | p | 1:32 | p | | |
| **CH43** | 1:3921 | p | 1:2600 | p | 1:1280 | p | | |
| **CH44** | 1:1971 | p | 1:1280 | p | 1:320 | p | | |
| **CH23** | 1:336 | p | 1:104 | p | 1:32 | p | | |
| **CH48** | 1:2002 | p | 1:3584 | p | 1:320 | p | | |
| **CH50** | 1:833 | p | 1:896 | p | 1:320 | p | | |
| **CH51** | 1:143 | p | 1:896 | p | 1:1280 | p | | |
| **CH52** | 1:7032 | p | 1:12800 | p | 1:1280 | p | | |

**Supplementary Table 1**. **TBEV WT SNT results using TBEV strain Hypr.** [1] *TBEV reporter virus particle seroneutralization test; results are given as IC99; the cutoff for a positive result was set at ≥1:40;* [2] *TBEV wild type seroneutralization test; results are given as IC99; the cutoff for a positive result was set at >1:8;* [3] *discordance of qualitative results between different tests are indicated with x.*

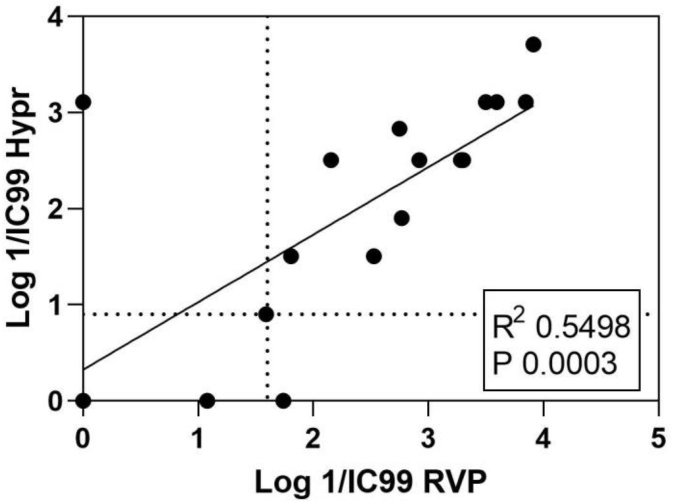


**Supplementary Figure 4. Linear regression analysis comparing results obtained using the TBEV RVP SNT and the WT SNT strain Hypr.** Results are expressed as log 1/IC99. Linear regression was calculated with GraphPad Prism, correlation coefficient and p values (T test for linear regression) are indicated in the panel. Dotted lines represent cut-off for qualitative analysis.

| | YFV RVP SNT | | YFV WT SNT | | Discording qualitative results |
|---|---|---|---|---|---|
| **sample** | **Ig tot. qn** | **Ig tot. ql** | **Ig tot. qn** | **Ig tot. ql** | **RVP SNT / WT SNT** |
| **CH41** | 1:13 | n | < 1:8 | n | |
| **CH13** | 1:22 | n | 1:8 | n | |
| **CH21** | 1:659 | p | 1:2048 | p | |
| **CH22** | 1:185 | p | 1:256 | p | |
| **CH23** | 1:1132 | p | 1:1024 | p | |
| **CH17** | 1:1588 | p | 1:2048 | p | |
| **CH40** | 1:1557 | p | 1:8192 | p | |
| **CH37** | 1:112 | p | 1:512 | p | |
| **CH39** | 1:90 | p | 1:256 | p | |
| **CH11** | 1:377 | p | 1:256 | p | |
| **CH15** | 1:1786 | p | 1:2048 | p | |

**Supplementary Table 2. Comparison of YFV RVP SNT and YFV WT SNT results.** [1] *YFV reporter virus particle seroneutralization test; results are given as IC99; the cutoff for a positive result was set at ≥1:40;* [2] *YFV wild type seroneutralization test; results are given as IC99; the cutoff for a positive result was set at >1:8;*
